# Apathy and Affective Symptoms Associated with Elevated Alzheimer’s Disease Biomarkers

**DOI:** 10.1101/2024.11.06.24316730

**Authors:** Matthew JY Kang, Dhamidhu Eratne, Samantha M Loi, Christa Dang, Alexander F Santillo, Henrik Zetterberg, Kaj Blennow, Philip B Mitchell, Malcolm Hopwood, Charles B Malpas, Dennis Velakoulis, Alzheimer’s Disease Neuroimaging Initiative

## Abstract

**Importance:** While apathy and affective neuropsychiatric symptoms (NPS) are common in the clinical spectrum of Alzheimer’s disease (AD), their neurobiological correlates remain unclear.

**Objective:** To longitudinally examine plasma markers of neurodegeneration (neurofilament light chain; NfL) and tau pathology (phosphorylated at threonine 181; p-tau181) and their associations with apathy and affective symptoms in individuals with clinical diagnoses of AD dementia and mild cognitive impairment (MCI).

**Design, Setting, Participants and Exposure:** This cohort study used longitudinal data from the Alzheimer Disease Neuroimaging Initiative (ADNI). Individuals with clinical diagnoses of MCI and AD dementia were enrolled in ADNI with serial annual blood samples over a four-year period, which were analyzed for NfL and p-tau181 using ultrasensitive techniques. Study data were accessed between July and September 2024.

**Main Outcomes and Measures:** The presence of neuropsychiatric symptoms was determined using the Neuropsychiatric Interview (NPI). We analyzed the trajectory of plasma NfL and p-tau181 in each NPS using general linear mixed-effects models adjusted for age and sex.

**Results:** There were 790 participants with AD dementia and MCI (mean [SD] age 72.7 [7.6] years; 333 females, 42%) and 417 healthy controls. The most common NPS was depression (n = 428; 54%), followed by irritability (n = 419; 53%), apathy (n = 348; 44%) and anxiety (n = 341; 43%). In the AD dementia and MCI group, apathy and anxiety were associated with higher levels of both NfL and p-tau181 after controlling for cognitive and functional decline. Moreover, apathy was associated with higher rate of NfL increase longitudinally. Depression at baseline was initially associated with higher NfL and p-tau181 levels, but this did not remain significant in the sensitivity analyses.

**Conclusion and Relevance:** This study found that in individuals with clinical AD, apathy and anxiety are associated greater tau and neurodegenerative burden. Furthermore, those with apathy had a higher rate of NfL increase, which suggests an accelerated neurodegenerative process. These findings highlight that these symptoms be indicative of AD pathology severity and have implications for potential treatments which target tau pathology.

**Key Points:** **Question**: Do apathy and affective neuropsychiatric symptoms correlate with the rate of tau pathology and neurodegeneration, as measured by plasma phosphorylated-tau181 (p-tau181) and neurofilament light chain protein (NfL), in individuals across the clinical spectrum of Alzheimer’s disease (AD)?

**Findings**: In a longitudinal study of 790 patients with mild cognitive impairment (MCI) and AD dementia, serial measurements of plasma p-tau181 and NfL were collected annually over four years. P-tau181 and NfL levels were elevated in those with apathy or anxiety compared to those without. The rate of NfL increase, an indicator of neurodegeneration, was significantly higher in those with apathy.

**Meaning**: The presence of anxiety and apathy was associated with elevated biomarkers of neurodegeneration in MCI and AD dementia. Notably, apathy was linked to a faster rate of NfL increase, suggesting an accelerated neurodegenerative process.

## Background

Neuropsychiatric symptoms (NPS) are prevalent in individuals across the clinical spectrum of Alzheimer’s disease (AD) including mild cognitive impairment (MCI) and AD dementia.^1,2^ These symptoms often co-occur in clusters, with affective symptoms (depression and anxiety) and apathy among the most prevalent.^1–4^ NPS have been associated with worse cognitive decline, higher mortality rates, and greater caregiver burden.^5–7^ Despite their prevalence and impact, the neurobiological mechanisms underlying NPS, and their contribution to disease progression remain inadequately understood.

Previous literature have reported mixed findings regarding the association between AD biomarkers and NPS.^8,9^ Several factors may account for the inconsistent results. Many studies may be underpowered to detect the subtle associations between NPS and biomarkers.^10^ Additionally, some studies have grouped NPS together,^2,6^ such as within the mild behavioral impairment (MBI) framework,^11,12^ potentially obscuring the effects of individual symptom groups.

Ultrasensitive blood-based immunoassays can now detect and track the progression of AD-related neuropathological processes. Plasma tau phosphorylated at threonine 181 (p-tau181) shows strong associations with tau pathology and amyloid deposition.^13^ Neurofilament light chain protein (NfL) reflects neuroaxonal damage,^14,15^ including in AD dementia.^16,17^ These blood-based biomarkers offer minimally invasive alternatives to traditional cerebrospinal fluid (CSF) and imaging biomarkers.^18,19^

A recent study found that that the emergence of psychosis was associated with elevated plasma levels of p-tau181 and NfL in individuals across the clinical spectrum of AD.^20^ However, no studies have explored their relationship to other NPS longitudinally, such as apathy and affective symptoms, which are the most common neuropsychiatric manifestations of AD dementia and MCI.

Our primary aim was to explore the association between apathy and affective NPS (depression, anxiety) with blood NfL and p-tau181 longitudinally in individuals with clinical diagnoses of AD dementia and MCI. As a secondary aim, we explored plasma biomarker associations with other NPS, including delusions, hallucinations, agitation, irritability, disinhibition, elation, appetite disturbance, sleep disturbance and aberrant motor symptoms.

## Methods

This longitudinal study used data from the Alzheimer’s Disease Neuroimaging Initiative (ADNI) database (adni.loni.usc.edu). The ADNI was launched in 2003 as a public-private partnership, led by Principal Investigator Michael W. Weiner, MD. The ADNI participants have been recruited from more than 50 sites across the United States and Canada. We downloaded the data from the ADNI database on the 22^nd^ July 2024. Regional ethical committees of all institutions approved the ADNI study. All study participants gave written informed consent. This study was carried out according to the Strengthening the Reporting of Observational Studies in Epidemiology (STROBE) reporting guideline.^21^

### Participants

This cohort has been described in a previous publication.^19^ In brief, the cohort consisted of patients with clinical diagnoses of AD dementia and MCI, and an additional group of cognitively unimpaired (CU) participants with available plasma NfL and p-tau181 measurements collected annually over four years. All ADNI participants had no significant neurological disease other than AD. The following measures were also obtained during the annual visits: Mini-Mental State Examination (MMSE), Clinical Dementia Rating Scale Sum of Boxes (CDR-SB), hippocampal volume acquired using 3-T MRI scanners,^22^ and Florbetapir positron emission tomography standardized uptake value ratio (AV45 PET SUVR) which quantifies amyloid plaque burden.^23^ The CU participants reported a MMSE score of 24 or higher and a CDR score of zero. The MCI participants reported an MMSE score of 24 or higher and a CDR score of 0.5. The patients with AD dementia fulfilled the clinical diagnostic criteria for probable AD,^24^ reported an MMSE score between 20 and 26, and a CDR score from 0.5 to 1.0.

The presence of NPS was assessed with the Neuropsychiatric Inventory (NPI), an informant-based scale that examines 12 neuropsychiatric domains through a structured interview with the caregiver. The NPS in the NPI are apathy, anxiety, depression, irritability, elation, disinhibition, aggression, delusions, hallucinations, aberrant motor symptoms, appetite disturbance and sleep disturbance.

For each NPS of interest (i.e. apathy), participants with AD dementia and MCI were distributed in 3 groups according to presence or absence of the NPS longitudinally: (1) participants without the specific NPS over the course of the study (i.e., apathy absent), (2) participants with the specific NPS at baseline (i.e., apathy baseline), and (3) participants with the specific NPS at any time point over the course of the study (i.e., apathy incident), consistent with a previous study by Gomar et al. exploring psychosis in the ADNI cohort.^20^

### Samples Analyses

Plasma p-tau181 and NfL concentrations were measured using Single molecule array (Simoa) technology (Quanterix, Billerica, MA) at the Clinical Neurochemistry Laboratory, University of Gothenburg, Mölndal, Sweden, as previously described.^19,20^

### Statistical Analyses

All statistical analyses were performed using R version 4.2.2 (2022-10-31), the lmer package, Jamovi version 2.6.0 and GAMLj package.^25–28^ For the participant characteristics, demographic and clinical status between groups at baseline were compared using general linear models (GLMs) for continuous variables, and χ^2^ test of independence were used for comparisons in categorical variables.

To explore whether a specific NPS (i.e. apathy) was associated with NfL or p-tau181 levels over time, a general linear mixed-effects model (GLMM) was estimated to analyze the longitudinal biomarkers (NfL and p-tau181) trajectories in each NPS subgroups (i.e. apathy absent, apathy baseline and apathy incident) with random intercepts and slopes for each participant. For the primary analysis, age and sex were included as covariates, using residual (restricted) maximum likelihood estimation and assuming an unstructured covariance matrix. Plasma NfL and p-tau181 levels were log-transformed. All continuous variables were centered and scaled prior to analysis. The rate of change of biomarkers within groups (β slopes) was computed as averaged individual slopes from GLMMs that included random intercepts and slopes. Robust standard errors and confidence intervals were computed using bootstrapping with 2000 replicates. We performed simple effects analysis to determine whether the presence of a specific NPS (i.e. apathy baseline vs apathy absent, apathy incident vs apathy absent) was associated with significantly higher NfL levels or slope compared to those without the specific NPS (i.e. absent baseline). All *p* values were 2-sided and corrected for multiple testing using the Benjamin-Hochberg adjusted false discovery rate (FDR) of 5%.

We performed additional **sensitivity analyses** to ensure the findings in the primary outcomes of interest (apathy, anxiety and depression) were robust. We added CDR-SB and MMSE scores as covariates to account for any confounding effects of dementia severity. We also repeated the GLMMs for the subgroup of individuals who demonstrated amyloid positivity during the study (AV45 SUVR > 1.11). As a post-hoc analysis, we also performed GLMMs using the severity score for each NPS derived from the NPI (frequency x severity) adjusting for age and sex as covariates.

## Results

A total of 1207 participants from ADNI had their serial plasma samples analyzed for p-tau181 and NfL are summarized in Table 1. For the cohort across the clinical AD spectrum (AD dementia and MCI; n = 790), the mean age was 72.7 (SD 7.63) years, with 333 females (42%). The mean CDR-SB was 5.3 for AD dementia and 1.5 for MCI, which is the equivalent of mild and very mild severity respectively. 161 (88%) individuals diagnosed with AD dementia and 327 (62%) individuals with MCI were amyloid-positive respectively. The most common NPS was depression (54%), followed by irritability (53%), apathy (44%) and anxiety (43%). The prevalence of the secondary NPS of interest are listed in sTable 1.

**Table 1-.**
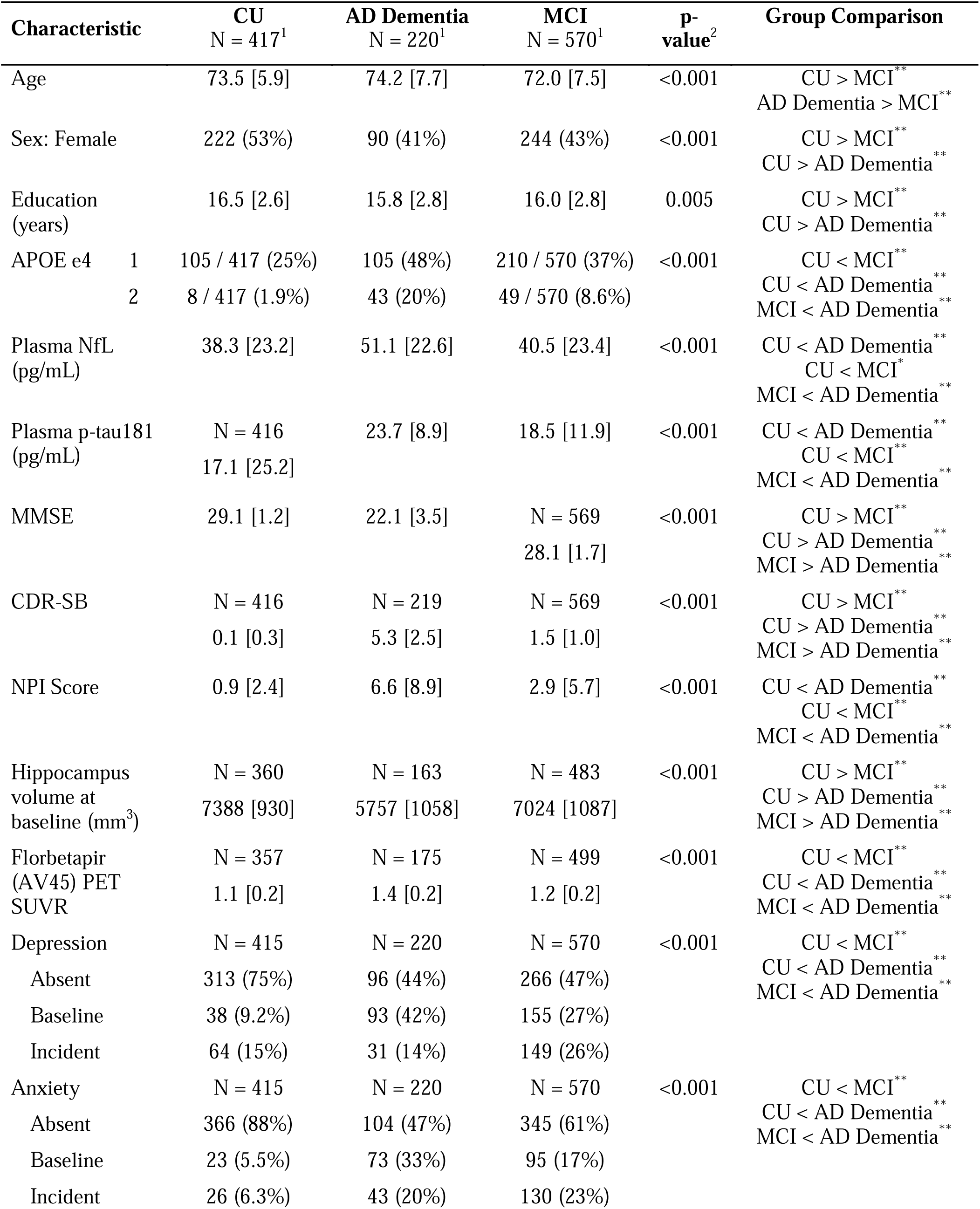
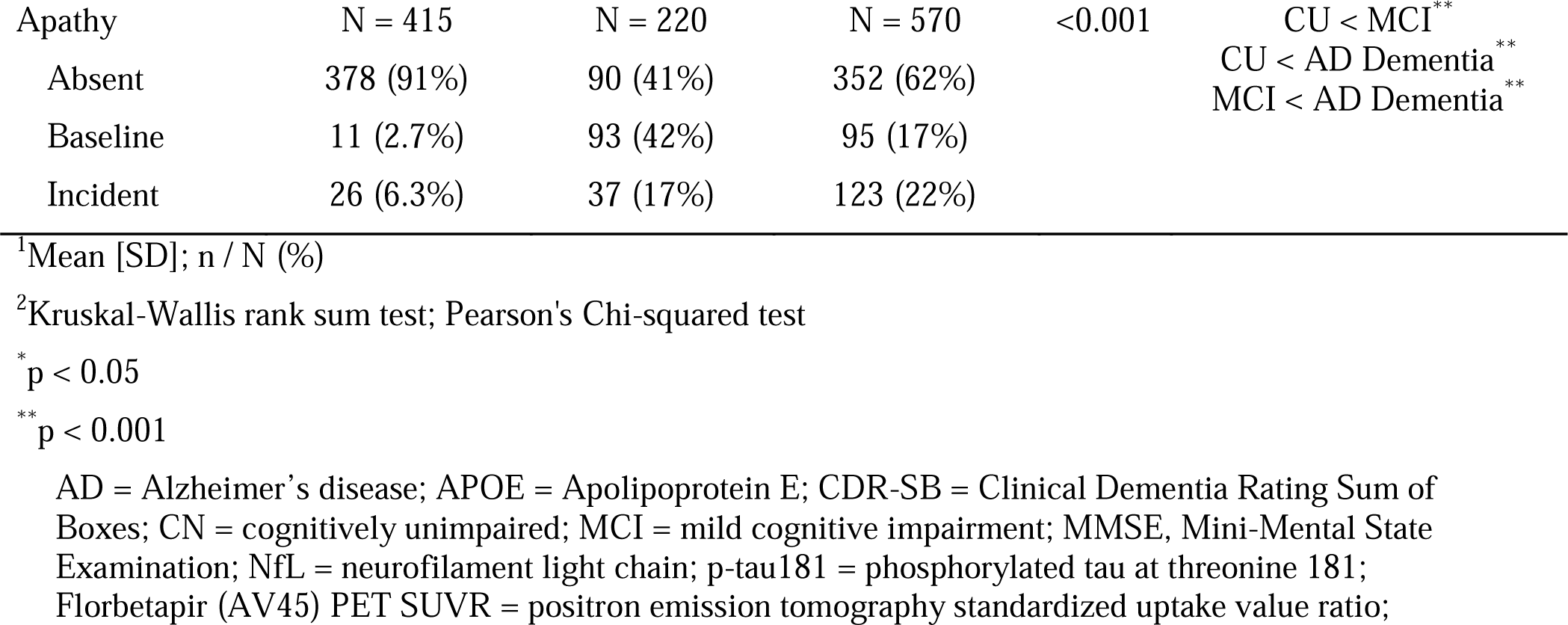
Participant Characteristics at Baseline.

### Associations between NPS and plasma NfL over time in the Clinical AD Spectrum

The overall associations between NPS and plasma NfL are shown in Figure 1. The primary NPS of interest are summarized in Figure 1 and Table 2.

**Figure 1-.**
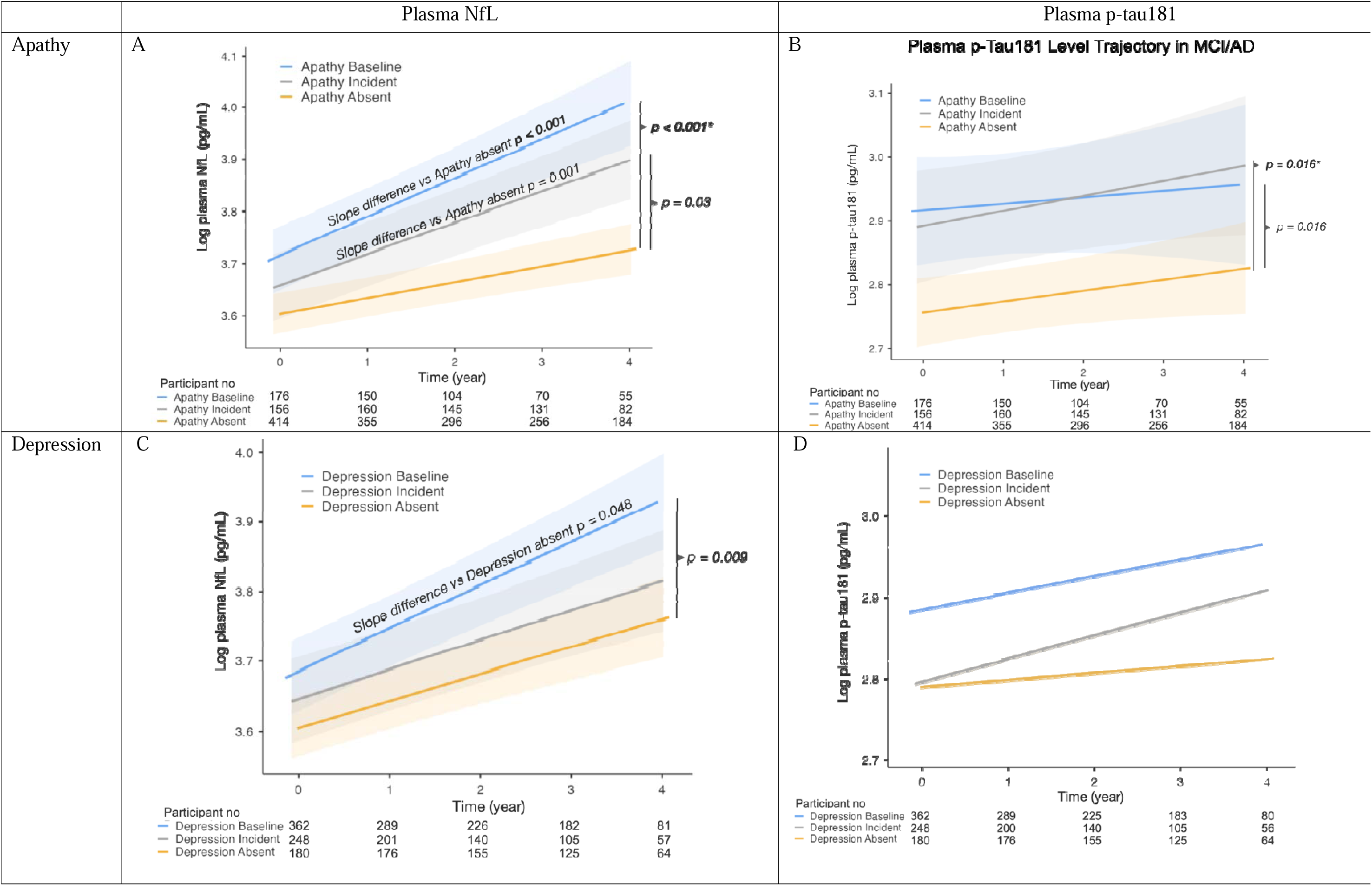
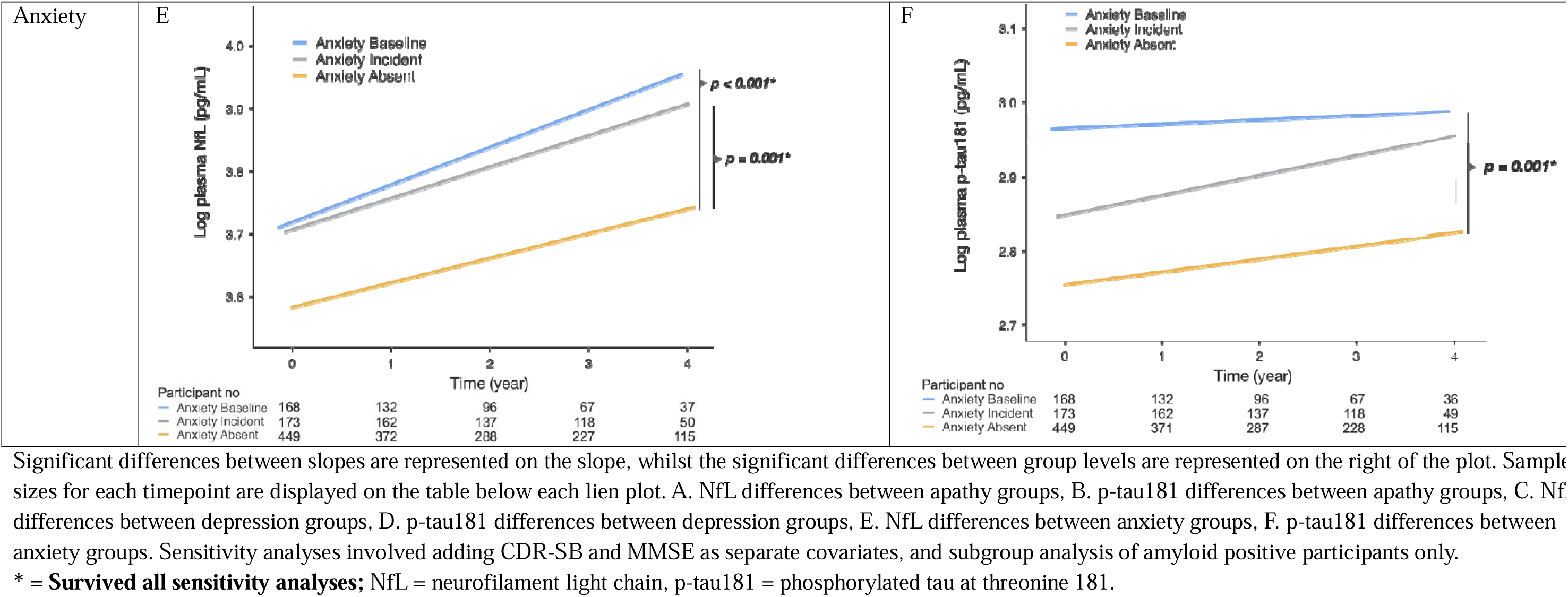
Longitudinal plots of plasma biomarkers for apathy, depression and anxiety in the clinical spectrum of Alzheimer’s disease

**Figure 2-.**
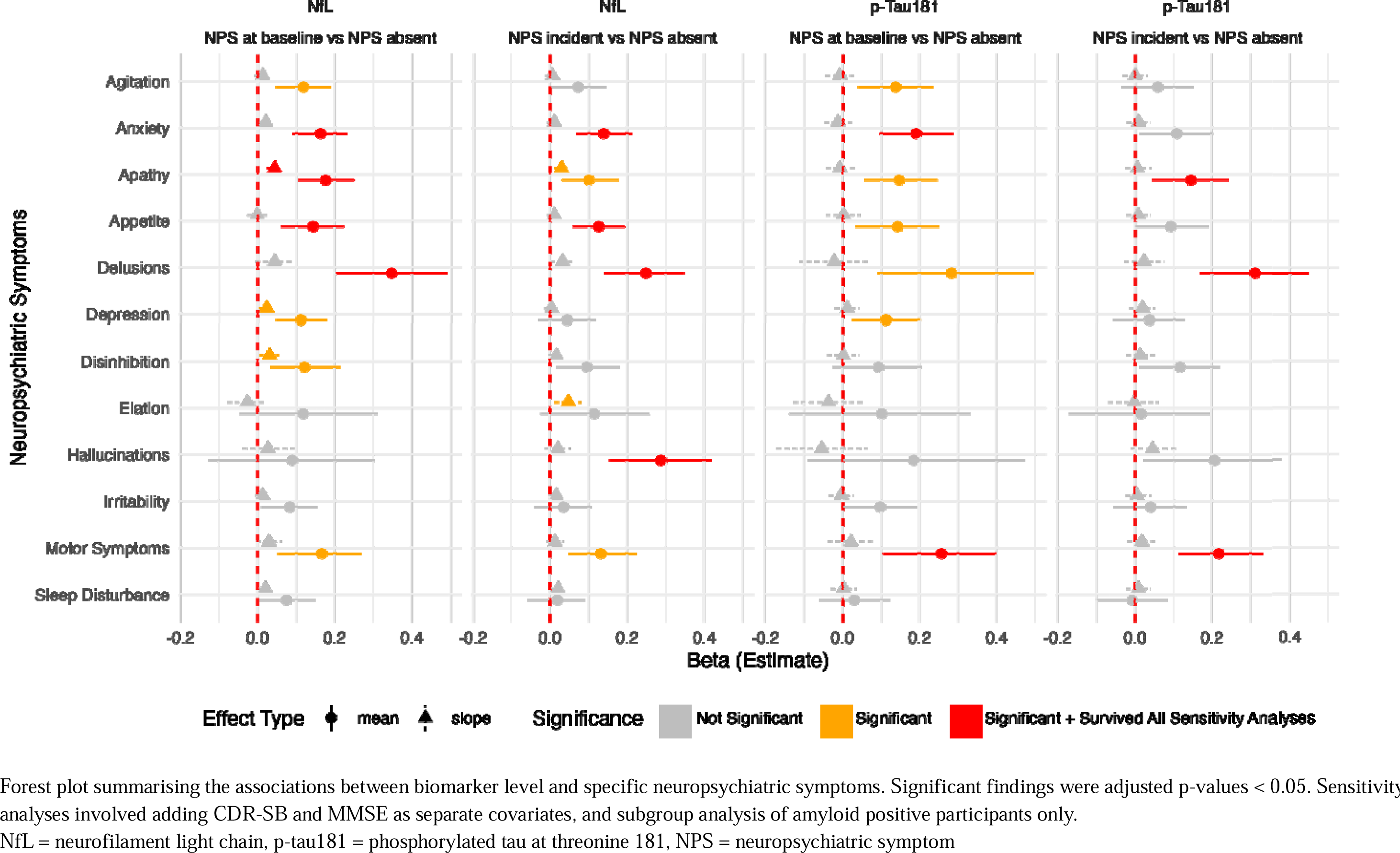
Forest plots of NPS effects on plasma biomarkers

**Table 2:** Summary of Apathy and Affective Symptom Effects on Plasma NfL and p-Tau181.

| NPS | Mean or Slope | Comparison Groups | Plasma NfL | Plasma p-tau181 |
| --- | --- | --- | --- | --- |
| Apathy | Mean | Baseline vs Absent | **0.18 (0.1, 0.25), p < 0.001<sup>s</sup>** | **0.15 (0.06, 0.24), p = 0.015** |
| Apathy | Mean | Incident vs Absent | **0.10 (0.03, 0.18), p = 0.029** | **0.14 (0.05, 0.24), p = 0.015<sup>s</sup>** |
| Apathy | Slope | Baseline vs Absent | **0.04 (0.02, 0.06), p < 0.001<sup>s</sup>** | -0.01 (-0.04, 0.03), p = 0.789 |
| Apathy | Slope | Incident vs Absent | **0.03 (0.01, 0.05), p = 0.009** | 0.01 (-0.03, 0.04), p = 0.789 |
| Depression | Mean | Baseline vs Absent | **0.11 (0.05, 0.18), p = 0.009** | **0.11 (0.02, 0.20), p = 0.048** |
| Depression | Mean | Incident vs Absent | 0.04 (-0.03, 0.12), p = 0.39 | 0.04 (-0.06, 0.13), p = 0.595 |
| Depression | Slope | Baseline vs Absent | **0.02 (0, 0.04), p = 0.048** | 0.01 (-0.02, 0.04), p = 0.623 |
| Depression | Slope | Incident vs Absent | 0 (-0.02, 0.02), p = 0.789 | 0.02 (-0.01, 0.05), p = 0.39 |
| Anxiety | Mean | Baseline vs Absent | **0.16 (0.09, 0.23), p < 0.001<sup>s</sup>** | **0.19 (0.1, 0.29), p = 0.001<sup>s</sup>** |
| Anxiety | Mean | Incident vs Absent | **0.14 (0.07, 0.21), p = 0.002<sup>s</sup>** | 0.11 (0.01, 0.20), p = 0.068 |
| Anxiety | Slope | Baseline vs Absent | 0.02 (0, 0.04), p = 0.118 | -0.01 (-0.05, 0.02), p = 0.665 |
| Anxiety | Slope | Incident vs Absent | 0.01 (-0.01, 0.03), p = 0.39 | 0.01 (-0.02, 0.04), p = 0.682 |

Note: Results are displayed as $\beta$ (95% confidence interval), p-value adjusted for multiple comparisons.
Analyses are corrected for age and gender.
**Bolded** indicates statistical significance set at $p < 0.05$
<sup>s</sup>Survived sensitivity analysis adding CDR-SB and MMSE as covariates separately
NfL = neurofilament light chain, p-tau181 = phosphorylated-tau 181, NPS = neuropsychiatric symptom

**Table 3:**
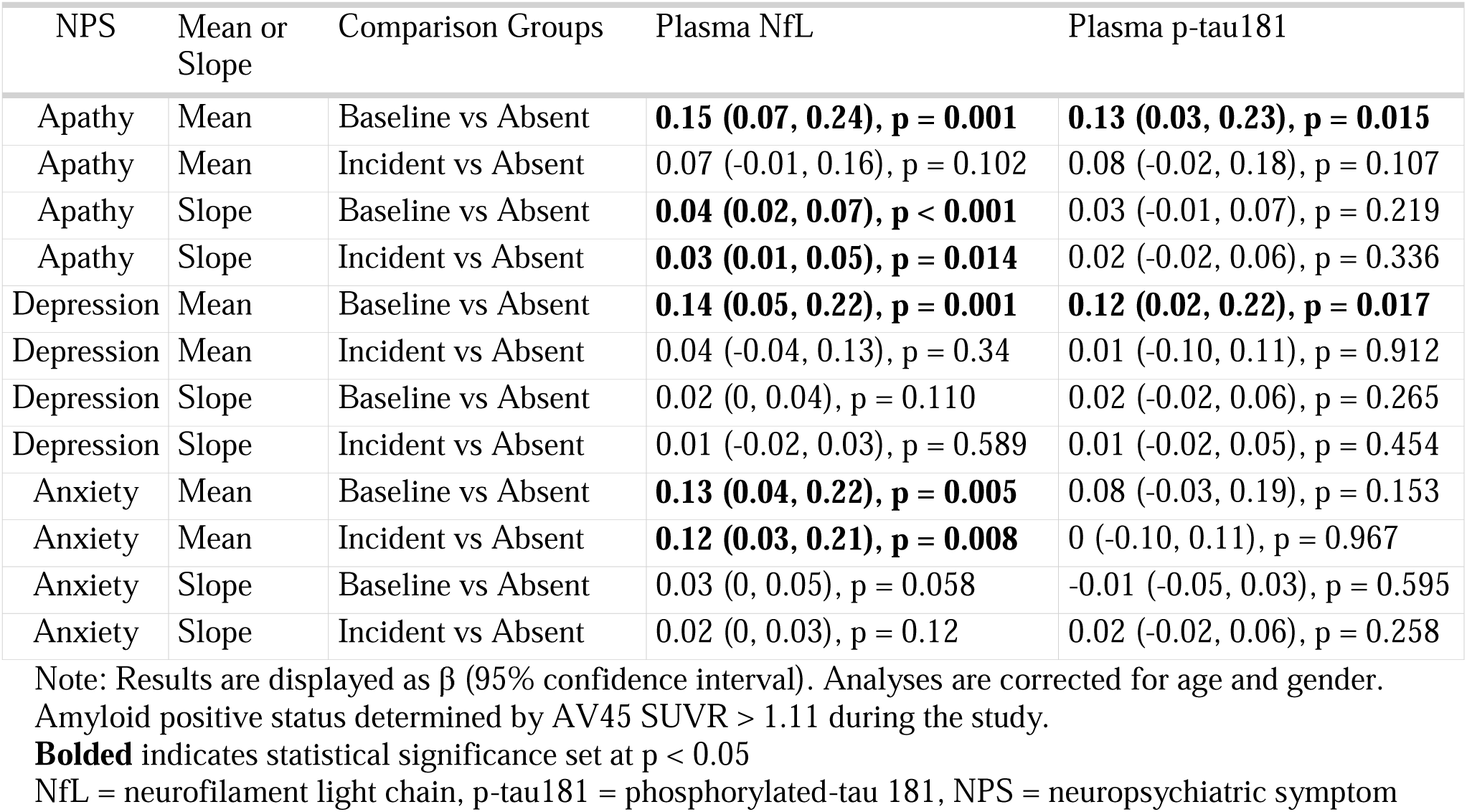
Subgroup analysis of Apathy and Affective Symptom Effects on Plasma NfL and p-Tau181 in amyloid positive group only.

#### Apathy

The GLMM revealed that the **apathy baseline group** had **higher NfL levels** over time, compared to the apathy absent group (β = 0.18, p < 0.001). This finding remained robust in sensitivity analyses adjusting for CDR-SB and MMSE. The **apathy incident group** had significantly higher NfL levels compared to the **apathy absent group** (β = 0.10, p = 0.029). This finding did not survive the sensitivity analyses. The simple effects analysis showed that both apathy baseline (β = 0.16, p < 0.001) and apathy incident (β = 0.09, p = 0.016 groups had higher NfL levels than the apathy absent group at the baseline timepoint.

In terms of the **rate of NfL increase, apathy baseline group** (β = 0.04, p < 0.001) **and apathy incident group** (β = 0.03, p = 0.009) exhibited higher rates of NfL increase compared to the **apathy absent group**. The significant finding for the apathy baseline group was robust in the sensitivity analyses.

#### Anxiety

The GLMM exploring the impact of anxiety on NfL levels showed that **anxiety absent group** had significantly higher NfL levels compared to the **anxiety absent group** (β = 0.16, p < 0.001). The **anxiety incident group** had significantly higher NfL levels than the **anxiety absent group** (β = 0.16, p < 0.001). Both findings were robust in the sensitivity analyses adjusting for CDR-SB and MMSE, and the subgroup analyses of amyloid positive individuals. The simple effects analysis also showed that both anxiety baseline (β = 0.16, p < 0.001) and anxiety incident (β = 0.14, p < 0.001) groups had higher NfL levels than the anxiety absent group at the baseline timepoint.

Neither anxiety baseline group nor anxiety incident group had significantly different rate of NfL increase compared to the anxiety absent group.

#### Depression

The GLMM examining depression found that the **depression baseline group** exhibited **significantly higher NfL levels** than the **depression absent group** (β = 0.11, p = 0.009). This finding was significant in the subgroup analysis of individuals who were amyloid positive, however this did not survive the sensitivity analyses adjusting for MMSE and CDR-SB. At the baseline timepoint, the depression baseline group had higher NfL levels than the depression absent group (β = 0.13; p < 0.001) whilst there were no significant differences between the depression incident and depression absent groups (β = 0.06, p = 0.104).

**The depression baseline group had a significantly higher rate of NfL increase** compared to the **depression absent group** (β = 0.02, p = 0.048). However, this did not survive the sensitivity analyses adjusting for CDR-SB and MMSE.

#### Other NPS of interest

For our secondary aims, the GLMMs found that baseline symptoms of delusions, appetite disturbances, agitation, disinhibition, and motor symptoms were associated with elevated NfL levels compared to those who did not develop the NPS over time. Individuals who developed (NPS incident) appetite disturbances, delusions, hallucinations, and motor symptoms were associated with increased NfL levels compared to their respective absent groups. Disinhibition baseline and elation incident showed significantly steeper increases in NfL levels compared to the absent groups.

#### Associations between each NPS severity score and NfL over time

For post-hoc analysis where each NPS was added to the GLMM as a severity variable, apathy and depression were associated with higher NfL levels, and the interaction terms for rate of increase were also significantly high. The post-hoc analyses are summarized in sTable 3.

### Associations between NPS and plasma p-tau181 over time in the Clinical AD Spectrum

The overall associations between NPS and plasma p-tau181 are shown in Figure 1. The primary NPS of interest are summarized in Figure 1 and Table 2. None of the NPS groups had significantly higher rates of p-tau181 change.

#### Apathy

GLMM showed that the apathy baseline group had higher p-tau181 levels compared to the apathy absent group (β = 0.15, p = 0.015). This finding was still significant in the subgroup analysis for amyloid positive individuals, but not in the sensitivity analyses controlling for CDR-SB. The apathy incident group also had higher p-tau181 levels compared to the apathy absent group (β = 0.14, p = 0.015). This finding survived sensitivity analyses controlling for CDR-SB and MMSE. At the baseline timepoint, both the apathy baseline (β = 0.24, p < 0.001) and apathy incident (β = 0.21, p < 0.001) groups had significantly higher levels compared to apathy absent group.

#### Depression

GLMM showed that the depression baseline group had higher p-tau181 levels compared to the depression absent group (β = 0.11, p = 0.048). This finding was still significant in the subgroup analysis of amyloid positive individuals but did not survive the sensitivity analyses adjusting for CDR-SB and MMSE. At the baseline timepoint, the depression baseline group had significantly higher levels compared to the depression absent group (β = 0.17, p < 0.001).

#### Anxiety

GLMM showed that the anxiety baseline group had higher p-tau181 levels compared to the anxiety absent group (β = 0.19, p = 0.001). This finding was robust in the sensitivity analyses controlling for CDR-SB and MMSE. At the baseline timepoint, both the anxiety baseline (β = 0.25, p < 0.001) and anxiety incident (β = 0.17, p = 0.001) groups had significantly higher levels compared to anxiety absent group.

#### Other NPS of interest

The secondary NPS of interest are summarized in sTable 2. Delusions, agitation, and motor symptoms at baseline were associated with higher p-tau181 levels compared to individuals without these symptoms. Furthermore, delusion incident and motor symptom incident also had higher p-tau181 levels compared to individuals without those symptoms.

#### Associations between each NPS severity score and NfL over time

When each NPS was added to the GLMM as a severity variable, depression severity was associated with p-tau181 levels (p = 0.009), whilst apathy severity was trending (p = 0.053).

## Discussion

In this longitudinal study, we found that plasma p-tau181 and NfL were significantly elevated in participants with apathy and anxiety at baseline or follow-up, compared to those without these symptoms. Moreover, participants with apathy showed a faster rate of NfL increase over time, suggesting accelerated neurodegeneration. Although depression showed an association with NfL and p-tau181, this did not remain significant in the sensitivity analyses controlling for cognitive and functional decline. These results underscore the potential relationship between neuropsychiatric symptoms and neurodegenerative biomarkers, particularly NfL, in the AD continuum.

Our findings support previous literature linking apathy with tau pathology and neurodegeneration.^29–32^ In particular, Kitamura et al. conducted one of the few multi-modal studies which investigated apathy in AD patients (n=17) with structural and tau imaging.^33^ They found an association between apathy, elevated tau deposition and decreased cortical thickness the in orbitofrontal cortex (OFC). The authors hypothesized that increased tau accumulation leads to degeneration of OFC thickness and integrity, thereby disrupting the limbic system, and contributing to apathy.^34,35^ Our observation that participants developing apathy also exhibited higher levels NfL and p-tau181 may be reflective of the preclinical manifestation of tau accumulation and neurodegeneration before the onset of clinical apathy. A longitudinal, multi-modal approach incorporating fluid-based AD biomarkers and neuroimaging is needed to further validate these findings.

Notably, some studies did not find an association between tau pathology and apathy.^29,36–39^ This discrepancy may be due to these studies being underpowered or cross-sectional,^37,40^ limiting their ability to detect subtle differences in tau pathology. Our study demonstrates the advantage of using blood-based AD biomarkers, which are more accessible and allow for larger, longitudinal data sets.^10,41^ However, a key limitation of blood biomarkers is that they provide a measure of overall pathology, without offering insights into specific brain regions or circuits directly involved.

The higher rate of NfL increase over time suggests heightened neurodegeneration in AD dementia and MCI patients with apathy, independent of cognitive and functional decline. Apathy has been linked to more rapid disease progression in AD, with several studies identifying it as a predictor of faster cognitive decline^42^ and earlier mortality.^43^ This may be due to the presence of apathy resulting in less optimal dementia care given its impact on functioning and caregiver burden, leading to worse outcomes.^42^ These findings reinforce the importance of identifying and treating neuropsychiatric symptoms including apathy in AD patients.^44^ Furthermore, plasma NfL may be a candidate for apathy-specific biomarker in AD to guide treatment.^45^

Anxiety was significantly associated with both NfL and p-tau181 in our study, aligning with previous reports linking anxiety to AD and neurodegenerative markers.^36,39^ Our longitudinal study builds on recent literature in CSF and cross-sectional data, including Banning et al’s investigation which found an association between anxiety and CSF p-tau181.^29^ Anxiety has been conceptualized as an early manifestation of compensatory behavior in response to preclinical cognitive decline,^29,46^ suggesting that its presence may serve as a behavioral signal of p-tau181 accumulation and subsequent neurodegeneration.

Although depression has been linked to AD progression,^47–49^ our study did not find a significant association between depression and NfL or p-tau181 after controlling for AD severity, consistent with other research.^29,36,50^ One hypothesis is that previous studies may have conflated measures of depression with other NPS, given the significant overlap between depression with apathy and anxiety. For example, Krell-Roesch et al. observed a relationship between clinical depression as defined by Beck Depression Inventory II (BDI-II) score and AD biomarkers, but not when depression was based on informant-observed NPI-Q.^9^ As several of the 21 items in BDI-II may be influenced by apathy, including loss of interest, energy and pleasure, the presence of apathy may have increased their BDI-II total score to meeting the cut-off score for depression. This highlights the heterogeneous nature of depression, and its apparent association with AD biomarkers may depend on the instrument that is used to define depression. However, our interpretation in this context is limited to depression symptoms, as our study could not control for a history of depression due to the lack of available data. Future studies that better characterize depression history in participants are needed.

As for our secondary aims, our study replicated findings that delusions and hallucinations are associated with higher NfL and p-tau181 levels.^20,51^ Our results also align with existing literature showing that agitation^50^ and motor symptoms^51^ are associated with disease progression.

There are several limitations of this study. The number of participants with available biomarker samples decreased during follow-up. As ADNI excluded participants with a history of major depression or bipolar disorder within the past year, it limits the generalizability of our findings to individuals with more severe psychiatric comorbidities, which is important given that mood disorders may have a mild influence on plasma NfL.^53^ Our study focused on p-tau181, but other tau species like p-tau217 may offer additional insights into tau pathology in AD. Future studies should explore the role of different tau species in relation to neuropsychiatric symptoms.

In conclusion, our findings emphasize the complex relationship between NPS and AD biomarkers, particularly NfL, and suggest that apathy may be linked to accelerated neurodegeneration. The observed changes in AD biomarkers across various NPS profiles provide valuable insights into the neurobiological mechanisms underlying these symptoms. Further research is required to better understand the role of these symptoms in disease progression and to develop targeted interventions aimed at slowing cognitive and functional decline in AD.

## Data Availability

All data produced are available online at https://adni.loni.usc.edu/

## CRediT authorship contribution statement

Matthew JY Kang: Study conceptualisation, literature review, data analysis, manuscript writing and submission

Dhamidhu Eratne: Study design, data analysis, manuscript revision and supervision

Samantha M Loi manuscript revision and supervision

Christa Dang manuscript revision

Alexander F Santillo manuscript revision and supervision

Henrik Zetterberg: Study design, biomarker data generation, manuscript revision

Kaj Blennow: Study design, biomarker data generation, manuscript revision

Philip B Mitchell: Study design, supervision and manuscript revision

Malcolm Hopwood: Study design, supervision and manuscript revision

Charles B Malpas: Study design, data analysis, statistical support, supervision and manuscript revision

Dennis Velakoulis: Study design, data analysis, supervision and manuscript revision

## Acknowledgements

Data collection and sharing for the Alzheimer’s Disease Neuroimaging Initiative (ADNI) is funded by the National Institute on Aging (National Institutes of Health Grant U19AG024904). The grantee organization is the Northern California Institute for Research and Education. In the past, ADNI has also received funding from the National Institute of Biomedical Imaging and Bioengineering, the Canadian Institutes of Health Research, and private sector contributions through the Foundation for the National Institutes of Health (FNIH) including generous contributions from the following: AbbVie, Alzheimer’s Association; Alzheimer’s Drug Discovery Foundation; Araclon Biotech; BioClinica, Inc.; Biogen; Bristol-Myers Squibb Company; CereSpir, Inc.; Cogstate; Eisai Inc.; Elan Pharmaceuticals, Inc.; Eli Lilly and Company; EuroImmun; F. Hoffmann-La Roche Ltd and its affiliated company Genentech, Inc.; Fujirebio; GE Healthcare; IXICO Ltd.; Janssen Alzheimer Immunotherapy Research & Development, LLC.; Johnson & Johnson Pharmaceutical Research & Development LLC.; Lumosity; Lundbeck; Merck & Co., Inc.; Meso Scale Diagnostics, LLC.; NeuroRx Research; Neurotrack Technologies; Novartis Pharmaceuticals Corporation; Pfizer Inc.; Piramal Imaging; Servier; Takeda Pharmaceutical Company; and Transition Therapeutics.

MK is supported by the Nick Christopher PhD scholarship and the Research Training Program Scholarship from the Department of Psychiatry, University of Melbourne with contributions from the Australian Commonwealth Government, and the Ramsay Hospital Research Foundation.

A.F.S is primarily funded by the Swedish federal government under the ALF agreement (ALF ALF 2022 YF 0017) The Fromma foundation and The Åke Wiberg foundation.

HZ is a Wallenberg Scholar and a Distinguished Professor at the Swedish Research Council supported by grants from the Swedish Research Council (#2023-00356; #2022-01018 and #2019-02397), the European Union’s Horizon Europe research and innovation programme under grant agreement No 101053962, Swedish State Support for Clinical Research (#ALFGBG-71320), the Alzheimer Drug Discovery Foundation (ADDF), USA (#201809-2016862), the AD Strategic Fund and the Alzheimer’s Association (#ADSF-21-831376-C, #ADSF-21-831381-C, #ADSF-21-831377-C, and #ADSF-24-1284328-C), the European Partnership on Metrology, co-financed from the European Union’s Horizon Europe Research and Innovation Programme and by the Participating States (NEuroBioStand, #22HLT07), the Bluefield Project, Cure Alzheimer’s Fund, the Olav Thon Foundation, the Erling-Persson Family Foundation, Familjen Rönströms Stiftelse, Stiftelsen för Gamla Tjänarinnor, Hjärnfonden, Sweden (#FO2022-0270), the European Union’s Horizon 2020 research and innovation programme under the Marie Skłodowska-Curie grant agreement No 860197 (MIRIADE), the European Union Joint Programme – Neurodegenerative Disease Research (JPND2021-00694), the National Institute for Health and Care Research University College London Hospitals Biomedical Research Centre, and the UK Dementia Research Institute at UCL (UKDRI-1003).

KB is supported by the Swedish Research Council (#2017-00915 and #2022-00732), the Swedish Alzheimer Foundation (#AF-930351, #AF-939721, #AF-968270, and #AF-994551), Hjärnfonden, Sweden (#FO2017-0243 and #ALZ2022-0006), the Swedish state under the agreement between the Swedish government and the County Councils, the ALF-agreement (#ALFGBG-715986 and #ALFGBG-965240), the European Union Joint Program for Neurodegenerative Disorders (JPND2019-466-236), the Alzheimer’s Association 2021 Zenith Award (ZEN-21-848495), the Alzheimer’s Association 2022-2025 Grant (SG-23-1038904 QC), La Fondation Recherche Alzheimer (FRA), Paris, France, the Kirsten and Freddy Johansen Foundation, Copenhagen, Denmark, and Familjen Rönströms Stiftelse, Stockholm, Sweden.

PBM is supported by an Australian NHMRC Investigator Grant (1177991), Lansdowne Foundation and Good Talk charity

## Data Sharing Statement

This study used data from the Alzheimer’s Disease Neuroimaging Initiative (ADNI; adni.loni.usc.edu)

## Conflicts of interest

HZ has served at scientific advisory boards and/or as a consultant for Abbvie, Acumen, Alector, Alzinova, ALZPath, Amylyx, Annexon, Apellis, Artery Therapeutics, AZTherapies, Cognito Therapeutics, CogRx, Denali, Eisai, LabCorp, Merry Life, Nervgen, Novo Nordisk, Optoceutics, Passage Bio, Pinteon Therapeutics, Prothena, Red Abbey Labs, reMYND, Roche, Samumed, Siemens Healthineers, Triplet Therapeutics, and Wave, has given lectures sponsored by Alzecure, BioArctic, Biogen, Cellectricon, Fujirebio, Lilly, Novo Nordisk, Roche, and WebMD, and is a co-founder of Brain Biomarker Solutions in Gothenburg AB (BBS), which is a part of the GU Ventures Incubator Program (outside submitted work).

KB has served as a consultant and at advisory boards for Abbvie, AC Immune, ALZPath, AriBio, BioArctic, Biogen, Eisai, Lilly, Moleac Pte. Ltd, Neurimmune, Novartis, Ono Pharma, Prothena, Roche Diagnostics, Sanofi and Siemens Healthineers; has served at data monitoring committees for Julius Clinical and Novartis; has given lectures, produced educational materials and participated in educational programs for AC Immune, Biogen, Celdara Medical, Eisai and Roche Diagnostics; and is a co-founder of Brain Biomarker Solutions in Gothenburg AB (BBS), which is a part of the GU Ventures Incubator Program, outside the work presented in this paper.

SL has received honorarium from Lundbeck and Otsuka.

MH has served as a consultant and at advisory boards for Boehringer-Ingelheim, Cortexa, Eli Lilly, Janssen, Lundbeck, Novartis, Otsuka, Orion, Servier and Takeda. He has received research support from Alto Neuroscience, Boehringer-Ingelheim, Douglas, Janssen, and Lyndra, PBM has received honoraria from Janssen (Australia) for lectures and advisory board membership

The remaining authors declare no conflict of interests.

## Notes

### Author Declarations

Ethical approval for the Alzheimer Disease Neuroimaging Initiative (ADNI) study was obtained from the medical ethics committees of all participating institutions, and written informed consent was obtained from all participants.

