## Supplemental Table for "Apathy and Affective Symptoms Associated with Elevated Alzheimer’s Disease Biomarkers"

sTable 1 – Prevalence of non-mood related neuropsychiatric symptoms

| **Characteristic** | **CU** | **Dementia** | **MCI** | **p-value^2^** | **Post-hoc Comparison** |
| --- | --- | --- | --- | --- | --- |
| Agitation | N = 415 | N = 220 | N = 570 | <0.001 | CU < MCI^**^ CU < Dementia^**^ MCI < Dementia^**^ |
| Absent | 367 (88%) | 112 (51%) | 341 (60%) |  |  |
| Baseline | 15 (3.6%) | 76 (35%) | 97 (17%) |  |  |
| Incident | 33 (8.0%) | 32 (15%) | 132 (23%) |  |  |
| Delusions | N = 415 | N = 220 | N = 570 | <0.001 | CU < MCI^**^ CU < Dementia^**^ MCI < Dementia^**^ |
| Absent | 409 (99%) | 164 (75%) | 531 (93%) |  |  |
| Baseline | 0 (0%) | 23 (10%) | 12 (2.1%) |  |  |
| Incident | 6 (1.4%) | 33 (15%) | 27 (4.7%) |  |  |
| Hallucinations | N = 415 | N = 220 | N = 570 | <0.001 | CU < MCI^**^ CU < Dementia^**^ MCI < Dementia^**^ |
| Absent | 412 (99%) | 191 (87%) | 543 (95%) |  |  |
| Baseline | 0 (0%) | 12 (5.5%) | 4 (0.7%) |  |  |
| Incident | 3 (0.7%) | 17 (7.7%) | 23 (4.0%) |  |  |
| Disinhibition | N = 415 | N = 220 | N = 570 | <0.001 | CU < MCI^**^ CU < Dementia^**^ MCI < Dementia^**^ |
| Absent | 383 (92%) | 140 (64%) | 427 (75%) |  |  |
| Baseline | 7 (1.7%) | 54 (25%) | 54 (9.5%) |  |  |
| Incident | 25 (6.0%) | 26 (12%) | 89 (16%) |  |  |
| Motor Symptom | N = 415 | N = 220 | N = 570 | <0.001 | CU < MCI^**^ CU < Dementia^**^ MCI < Dementia^**^ |
| Absent | 409 (99%) | 148 (67%) | 482 (85%) |  |  |
| Baseline | 3 (0.7%) | 37 (17%) | 25 (4.4%) |  |  |
| Incident | 3 (0.7%) | 35 (16%) | 63 (11%) |  |  |
| Sleep Symptoms | N = 415 | N = 220 | N = 568 | <0.001 | CU < MCI^**^ CU < Dementia^**^ |
| Absent | 283 (68%) | 141 (64%) | 298 (52%) |  |  |
| Baseline | 52 (13%) | 53 (24%) | 128 (23%) |  |  |
| Incident | 80 (19%) | 26 (12%) | 142 (25%) |  |  |
| Appetite Symptoms | N = 414 | N = 220 | N = 570 | <0.001 | CU < MCI^**^ CU < Dementia^**^ MCI < Dementia^**^ |
| Absent | 364 (88%) | 123 / 220 (56%) | 393 (69%) |  |  |
| Baseline | 12 (2.9%) | 62 / 220 (28%) | 54 (9.5%) |  |  |
| Incident | 38 (9.2%) | 35 / 220 (16%) | 123 (22%) |  |  |
| Elation | N = 415 | N = 220 | N = 570 | <0.001 | CU < MCI^**^ CU < Dementia^**^ |
| Absent | 410 (99%) | 206 (94%) | 530 (93%) |  |  |
| Baseline | 0 (0%) | 8 (3.6%) | 14 (2.5%) |  |  |
| Incident | 5 (1.2%) | 6 (2.7%) | 26 (4.6%) |  |  |
| Irritability | N = 415 | N = 220 | N = 570 | <0.001 | CU < MCI^**^ CU < Dementia^**^ MCI < Dementia^**^ |
| Absent | 314 (76%) | 106 (48%) | 265 (46%) |  |  |
| Baseline | 43 (10%) | 80 (36%) | 145 (25%) |  |  |
| Incident | 58 (14%) | 34 (15%) | 160 (28%) |  |  |
| ^1^Mean [SD]; n / N (%) | | | | | |
| ^2^Kruskal-Wallis rank sum test; Pearson's Chi-squared test  ^*^p < 0.05  ^**^p < 0.001  CU = cognitively unimpaired; MCI = mild cognitive impairment | | | | | |

sTable 2: Summary of NPS Effects on Plasma NfL and p-Tau181

| NPS | Mean or Slope | Comparison Groups | Plasma NfL | Plasma p-Tau181 |
| --- | --- | --- | --- | --- |
| Delusions | Mean | Baseline vs absent | **0.35 (0.2, 0.49),** **p < 0.001^s^** | **0.28 (0.09, 0.49), p = 0.026** |
| Delusions | Mean | Incident vs absent | **0.25 (0.14, 0.35), p < 0.001^s^** | **0.31 (0.17, 0.45), p < 0.001^s^** |
| Delusions | Slope | Baseline vs absent | 0.04 (-0.01, 0.1), p = 0.221 | -0.02 (-0.11, 0.07), p = 0.755 |
| Delusions | Slope | Incident vs absent | 0.03 (0, 0.06), p = 0.089 | 0.02 (-0.03, 0.08), p = 0.559 |
| Hallucinations | Mean | Baseline vs absent | 0.09 (-0.13, 0.3), p = 0.576 | 0.19 (-0.09, 0.47), p = 0.384 |
| Hallucinations | Mean | Incident vs absent | **0.29 (0.15, 0.42), p < 0.001^s^** | 0.21 (0.02, 0.38), p = 0.055 |
| Hallucinations | Slope | Baseline vs absent | 0.03 (-0.04, 0.1), p = 0.595 | -0.05 (-0.17, 0.06), p = 0.543 |
| Hallucinations | Slope | Incident vs absent | 0.02 (-0.01, 0.05), p = 0.39 | 0.05 (-0.01, 0.1), p = 0.249 |
| Agitation | Mean | Baseline vs absent | **0.12 (0.05, 0.19), p = 0.01** | **0.14 (0.04, 0.24), p = 0.026** |
| Agitation | Mean | Incident vs absent | 0.07 (0, 0.14), p = 0.125 | 0.06 (-0.03, 0.15), p = 0.39 |
| Agitation | Slope | Baseline vs absent | 0.01 (-0.01, 0.03), p = 0.39 | -0.01 (-0.04, 0.03), p = 0.789 |
| Agitation | Slope | Incident vs absent | 0.01 (-0.01, 0.02), p = 0.623 | 0 (-0.03, 0.03), p = 0.985 |
| Disinhibition | Mean | Baseline vs absent | 0.12 (0.03, 0.21), p = 0.026 | 0.09 (-0.03, 0.2), p = 0.223 |
| Disinhibition | Mean | Incident vs absent | 0.1 (0.02, 0.18), p = 0.068 | 0.12 (0.01, 0.22), p = 0.088 |
| Disinhibition | Slope | Baseline vs absent | **0.03 (0.01, 0.05), p = 0.048** | 0 (-0.04, 0.04), p = 0.944 |
| Disinhibition | Slope | Incident vs absent | 0.02 (0, 0.04), p = 0.223 | 0.01 (-0.02, 0.05), p = 0.595 |
| Motor Symptoms | Mean | Baseline vs absent | **0.17 (0.05, 0.27), p = 0.015** | **0.26 (0.11, 0.4), p = 0.004^s^** |
| Motor Symptoms | Mean | Incident vs absent | **0.13 (0.05, 0.22), p = 0.015** | **0.22 (0.11, 0.33), p = 0.002^s^** |
| Motor Symptoms | Slope | Baseline vs absent | 0.03 (0, 0.06), p = 0.202 | 0.02 (-0.04, 0.08), p = 0.601 |
| Motor Symptoms | Slope | Incident vs absent | 0.01 (-0.01, 0.03), p = 0.401 | 0.02 (-0.02, 0.06), p = 0.543 |
| Sleep Disturbance | Mean | Baseline vs absent | 0.07 (0.01, 0.15), p = 0.103 | 0.03 (-0.06, 0.12), p = 0.665 |
| Sleep Disturbance | Mean | Incident vs absent | 0.02 (-0.06, 0.09), p = 0.721 | -0.01 (-0.1, 0.08), p = 0.928 |
| Sleep Disturbance | Slope | Baseline vs absent | 0.02 (0, 0.04), p = 0.088 | 0 (-0.03, 0.04), p = 0.924 |
| Sleep Disturbance | Slope | Incident vs absent | 0.02 (0, 0.04), p = 0.068 | 0.01 (-0.02, 0.04), p = 0.682 |
| Appetite | Mean | Baseline vs absent | **0.14 (0.06, 0.22), p = 0.004****^s^** | 0.14 (0.04, 0.25), p = 0.048 |
| Appetite | Mean | Incident vs absent | **0.13 (0.06, 0.19), p = 0.002^s^** | 0.09 (0, 0.19), p = 0.125 |
| Appetite | Slope | Baseline vs absent | 0 (-0.03, 0.02), p = 0.928 | 0 (-0.04, 0.05), p = 0.948 |
| Appetite | Slope | Incident vs absent | 0.01 (-0.01, 0.03), p = 0.39 | 0.01 (-0.02, 0.04), p = 0.707 |
| Elation | Mean | Baseline vs Absent | 0.12 (-0.05, 0.31), p = 0.367 | 0.1 (-0.14, 0.33), p = 0.571 |
| Elation | Mean | Incident vs Absent | 0.11 (-0.02, 0.26), p = 0.24 | 0.02 (-0.17, 0.20), p = 0.924 |
| Elation | Slope | Baseline vs Absent | -0.03 (-0.08, 0.03), p = 0.5 | -0.04 (-0.13, 0.05), p = 0.595 |
| Elation | Slope | Incident vs Absent | **0.05 (0.01, 0.08), p = 0.038** | 0 (-0.07, 0.06), p = 0.975 |
| Irritability | Mean | Baseline vs Absent | 0.08 (0.01, 0.15), p = 0.06 | 0.1 (0, 0.2), p = 0.093 |
| Irritability | Mean | Incident vs Absent | 0.03 (-0.04, 0.11), p = 0.538 | 0.04 (-0.05, 0.13), p = 0.571 |
| Irritability | Slope | Baseline vs Absent | 0.01 (-0.01, 0.03), p = 0.292 | 0 (-0.04, 0.03), p = 0.924 |
| Irritability | Slope | Incident vs Absent | 0.02 (0, 0.03), p = 0.154 | 0.01 (-0.02, 0.04), p = 0.789 |

Note: Results are displayed as β (95% confidence interval), p-value adjusted for multiple comparisons. Analyses are corrected for age and gender.
**Bolded** indicates statistical significance set at p < 0.05
 ^*^Survived sensitivity analysis adding CDR-SB as a covariate
NfL = neurofilament light chain, p-tau181 = phosphorylated-tau 181, NPS = neuropsychiatric symptom

sTable 3: Post-hoc analyses of Apathy and Affective Symptom Severity’s Effects on Plasma NfL and p-Tau181

| NPS | Mean or Slope | Plasma NfL | Plasma p-tau181 |
| --- | --- | --- | --- |
| Apathy | Mean | **0.011 (0.005, 0.017), p < 0.001** | 0.010 (0, 0.020), p = 0.053 |
| Apathy | Slope | **0.007 (0.003, 0.011), p = 0.002** | 0.002 (-0.006, 0.009), p = 0.654 |
| Depression | Mean | **0.012 (0.004, 0.021), p = 0.003** | **0.018 (0.004, 0.032), p = 0.009** |
| Depression | Slope | **0.007 (0.001, 0.013), p = 0.014** | 0.002 (-0.008, 0.012), p = 0.652 |
| Anxiety | Mean | 0.006 (0.001, 0.014), p = 0.125 | **0.014 (0.001, 0.027), p = 0.036** |
| Anxiety | Slope | 0 (-0.006, 0.005), p = 0.817 | -0.009 (-0.018, 0.001), p = 0.070 |

Note: Results are displayed as β (95% confidence interval. Analyses are corrected for age and gender.
**Bolded** indicates statistical significance set at p < 0.05
NfL = neurofilament light chain, p-tau181 = phosphorylated-tau 181, NPS = neuropsychiatric symptom
